# Gait age clocks in health and disease

**DOI:** 10.64898/2026.08.14.26357566

**Authors:** Carlos Coronel-Oliveros, Fernando Lehue, Isabelle Killane, Joseph Mc Donnell, Silvin Knight, Mikel Gainza

## Abstract

Gait is a scalable biomarker of functional, physical, and brain health, but most studies rely on gait speed alone. Here, we developed and validated gait age clocks that estimate age from multidimensional gait features and quantify deviations as gait age gaps, with gaps >0 (<0) for accelerated (delayed) aging. We included data from 5,681 participants, including healthy controls and clinical groups (Parkinson’s disease, neurodegenerative diseases, stroke, diabetes, fallers, and frailty). Normative models trained in healthy controls showed robust age prediction (*r*=0.851, *p*<0.001), and full gait models outperformed gait speed alone (ΔR^2^=0.175). Gaps captured accelerated aging across neurological and physical conditions, tracked Parkinson’s disease severity, and were associated with frailty, physical performance, white matter hyperintensities, and geriatric depression. Gait age gaps are also related to brain aging, risk/protective lifestyle factors, and mortality risk. These findings support gait age gaps as an interpretable biomarker for aging, risk stratification, and clinical monitoring.

## Introduction

Gait speed is one of the most widely used biomarkers of functional ability, physical health, and brain health across aging and disease^1–3^. Gait assessment is scalable, affordable, and easy to incorporate into clinical routine examinations. Although useful, the complexity of gait dynamics contains more information than gait speed alone^3,4^. Features such as stride length, asymmetry, and gait balance capture additional aspects of motor and neural function and are altered in conditions such as Parkinson’s disease, frailty, and fall risk^4–6^. Normative modelling (clock models) can summarize this multidimensional information into a single gait health score by estimating age-related gait patterns and quantifying deviations from them^7–9^. Health scores from gait might be a promising alternative in clinical settings (compared to routine clinical examinations), considering the easy deployability of mobile phones and wearables as instruments for gait assessment^10^. However, gait clock models have not been robustly characterised using large samples across healthy and clinical populations.

Gait speed has been widely used in geriatrics and neurology as a reliable indicator of functional status and clinical risk^2–4^. It is routinely used to assess frailty, predict falls, and monitor mobility decline, and it is useful for tracking motor symptom progression in Parkinson’s disease^4^. Gait speed has also been linked to brain health in healthy aging and neurological disease, supporting the view that walking performance reflects central nervous system integrity in addition to peripheral motor function^2,11^. Still, gait speed captures only one aspect of gait. Other variables provide complementary information, including double support time (the proportion of the gait cycle during which both feet are simultaneously in contact with the ground) and walking asymmetry (differences between left and right step timing or step characteristics). Integrating these features may therefore provide a more informative description of health status than gait speed alone^4,12,13^. This is especially relevant now that mobile phones and wearables can capture gait with increasing quality in clinics and real-world settings^10,14,15^. Summarising multidimensional gait features into easy-to-measure, interpretable scores could make gait assessment a scalable and affordable tool for personalised medicine and tailored interventions.

Normative age models extend the idea of normative clinical data, but instead of providing reference ranges for single measures, they learn the multivariate pattern of features expected at each age in healthy individuals^16,17^. These data-driven models are trained in healthy individuals to learn the expected relationship between biological or behavioural features and chronological age^17^. These normative models predict participants’ age from the input features, and the difference between the predicted age and chronological age is called the “age gap”^9,17^. A positive age gap (gap > 0) indicates accelerated aging (the participant appears older than expected), whereas a negative one (gap < 0) indicates delayed aging^9,17^. Clock models have been developed for multiple domains, including organs’^9^, brain imaging^8,9,18^, proteomic markers^19^, and biobehavioural phenotypes^7^. They consistently show accelerated aging in dementia, neurological, and neuropsychiatric conditions^8,9,18^. They have also been used to characterize protective factors such as exercise, lifestyle, socioeconomic disparities, and creativity^7,9,20,21^. Despite the growth of this literature, gait clocks remain underexplored, and, to our knowledge, no gait clock model has been robustly validated in a large cohort using a broad set of gait parameters and clinically diverse samples. Given the low cost and minimal burden of gait analysis, gait age models could serve as a useful biomarker for preventive care, disease monitoring, and personalised medicine.

Here, we developed and validated a gait clock model in a large cohort of participants (N = 5,681), including healthy adults and people with Parkinson’s disease, other neurodegenerative diseases (Huntington’s disease and amyotrophic lateral sclerosis), diabetes, frailty, and a history of falls, with cross-sectional and longitudinal data. We used machine learning regression models with nested cross-validation to predict age from gait features and sex, and from the models, we computed the gait age and gaps^9,17^. We then compared gait age gaps across clinical groups and controls, tested whether gaps capture accelerated aging in clinical conditions, tracked disease severity and progression, and examined associations with biomarkers, lifestyle factors, MRI-based brain clocks, and mortality risk. We hypothesised that (i) full gait models using multidimensional gait dynamics would outperform gait speed-only models, (ii) gaps would capture accelerated aging in disease and reflect known associations with health biomarkers and lifestyle, and (iii) gaps would outperform individual gait parameters as biomarkers of mortality.

## Results

The overall pipeline is shown in **Fig. 1**. We pooled data from five open-source databases^22–28^ (open dataset, N = 562), and The Irish Longitudinal Study on Ageing study^29^ (TILDA dataset, N = 5,119), for a total sample of N = 5,681 participants (**Fig. 1A**, full demographics in **Table 1**; dataset details in **Supplementary Table S1**). Across the pooled sample, participants included healthy controls (N = 1,391) and non-healthy controls (N = 4,290). From this last subset, we included participants with Parkinson’s disease (PD, N = 120), neurodegenerative diseases (ND; Huntington’s disease and amyotrophic lateral sclerosis, N = 24), stroke (ST, N = 60), diabetes (N = 324), fallers (N = 86), and frailty (N = 49). The remaining non-HCs, although they were not used in group comparisons, were included in analyses of associations with risk/protective factors and mortality. Separate normative models of aging were trained, within each dataset, using HCs only (open dataset: N = 311, age range 40-97 years, 46% females; TILDA dataset: N = 1,080, age range 34-87 years, 54% females) and gait features plus biological sex. In the open dataset, model inputs included four gait parameters (speed, cadence, stride length, and double support time; **Fig. 1B**). In TILDA, models used 39 gait parameters (**Supplementary Table S2**). We designed a machine learning pipeline that included data augmentation (open dataset only), nested cross-validation, and support vector regression (**Fig. 1C**)^18^. Model predictions (“gait age”) were used to compute gaps (gaps = predicted minus chronological age), where gaps > 0 indicate accelerated aging and gaps < 0 indicate delayed aging^9^ (**Fig. 1D**).

**Figure 1.**
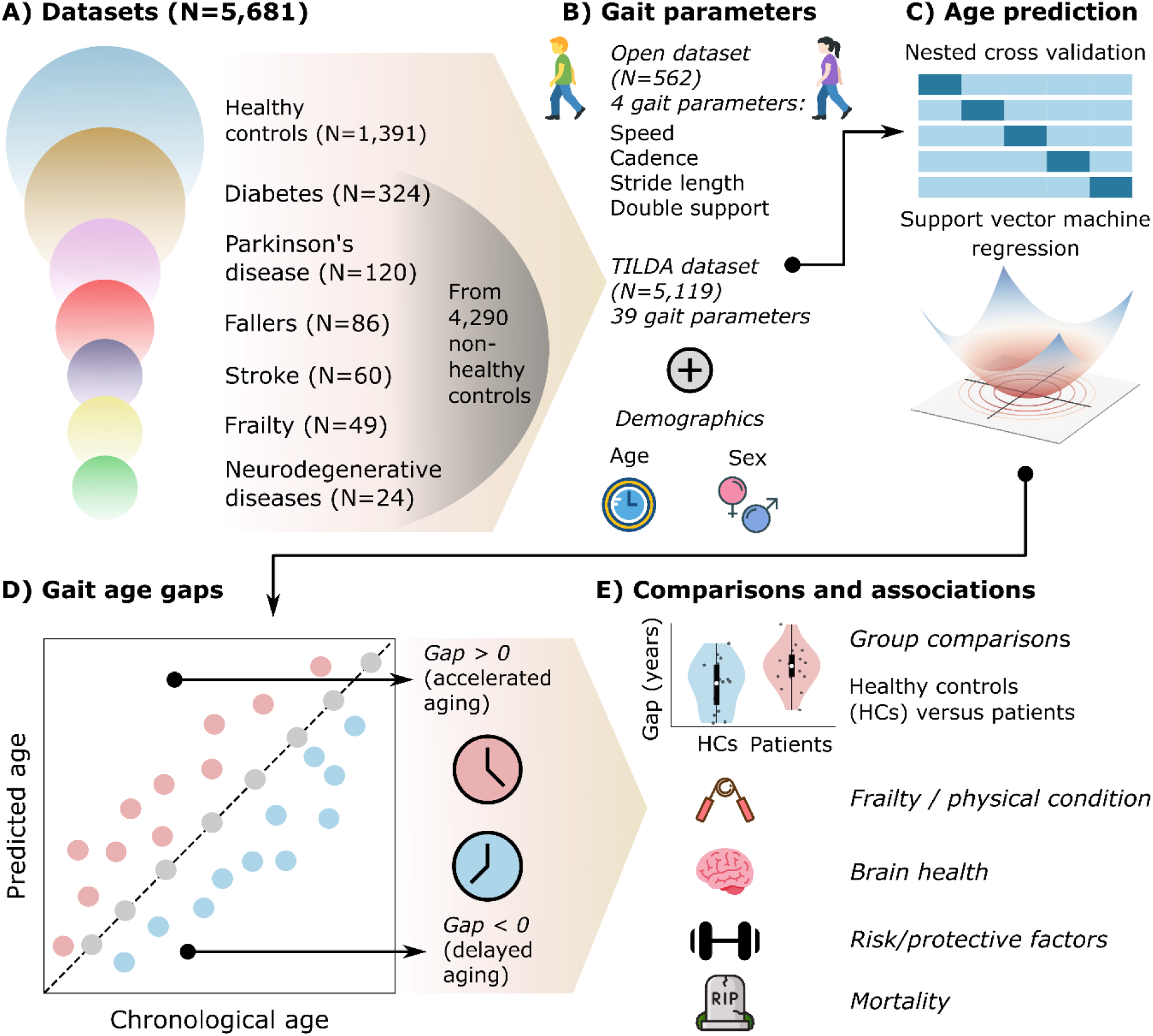
Participants and analytical pipeline. **A)** We included data from N=5,681 participants, comprising healthy controls (HCs, N=1,391), patients with diabetes (N=324), Parkinson’s disease (N=120, fallers (N=86), stroke (N=60), frailty (N=49), and neurodegenerative diseases (Huntington’s disease and amyotrophic lateral sclerosis, N=24). The patients were selected from a bigger pool of non-healthy controls (N=4,290). **B)** From open-source databases, we retrieved four gait parameters (speed, cadence, stride length, and double support time, among others), plus demographic and clinical/biological data (when available). **C)** The normative models of aging were trained using healthy controls’ gait parameters and biological sex. We used a computational pipeline comprising data augmentation (up to 50% new observations), nested cross-validation, and support vector machine regression. **D)** We assessed the models’ performance with Pearson’s correlation and mean absolute error between chronological age and predicted age (the “gait age”). Based on the predictions, we computed the gaps as the difference between predicted and chronological age. The gaps served as general health scores, with gap > 0 indicating accelerated aging and gap < 0 indicating delayed aging. **E)** Finally, we compared the gaps between HCs and the different clinical populations (patients), associated the gaps with physical and brain health markers as well as lifestyle factors, and finally, we used the gaps to predict mortality from longitudinal data.

**Table 1.** Participants and demographics.

| Open dataset |  |  |  |  |  |  |  |  |
| --- | --- | --- | --- | --- | --- | --- | --- | --- |
| Variables | Ful sample | HCs | PD | ND<br>(HT+AL) | Diabetes | Fallers | Anova / X<sup>2</sup>:<br>PD and ND vs HCs | Anova / X<sup>2</sup>:<br>Diabetes and<br>Fallers vs HCs |
| Sample size | 562 | 311 | 105 | 24 | 36 | 86 |  |  |
| Sex (F:M) | 298:264 | 166:145 | 38:67 | 12:12 | 15:21 | 67:19 | X<sup>2</sup> = 9.29<br>*p* = 0.010 | X<sup>2</sup> = 20.48<br>*p* < 0.001 |
| Age (years); range, mean ± standard deviation | (40-97)<br>64.86 ± 14.97 | (40-97)<br>59.56 ± 14.44 | (44-84)<br>66.49 ± 9.19 | (40-71)<br>54.58 ± 10.91 | (51-80)<br>66.33 ± 8.11 | (72-97)<br>84.33 ± 5.98 | Welch's F = 22.24<br>*p* < 0.001 | Welch's F = 298.09<br>*p* < 0.001 |
| TILDA dataset |  |  |  |  |  |  |  |  |
| Variables | Ful sample | HCs | PD | ST | Diabetes | Frailty | Anova / X<sup>2</sup>:<br>PD and ST vs HCs | Anova / X<sup>2</sup>:<br>Diabetes and<br>Frailty vs HCs |
| Sample size | 5,119 | 1,080 | 15 | 60 | 288 | 49 |  |  |
| Sex (F:M) | 2,858:2,261 | 585:495 | 7:8 | 30:30 | 110:178 | 28:21 | X<sup>2</sup> = 0.713<br>*p* = 0.700 | X<sup>2</sup> = 23.972<br>*p* < 0.001 |
| Age (years); range, mean ± standard deviation | (29-93)<br>61.10 ± 8.84 | (34-87)<br>57.26 ± 7.90 | (55-80)<br>67.20 ± 8.26 | (32-91)<br>67.03 ± 9.80 | (38-89)<br>64.58 ± 8.69 | (46-89)<br>65.84 ± 11.72 | Welch's F = 37.985<br>*p* < 0.001 | Welch's F = 92.491<br>*p* < 0.001 |

### Full gait models outperformed gait speed

We assessed model performance using Pearson’s correlation and mean absolute error (MAE) between chronological age and predicted age (averaged across folds and repetitions). The age bias correction was used to correct gaps, but it was not applied when reporting the model performance^30^. The full gait model yielded robust age estimation performance (*r* = 0.851, *p* < 0.001, R^2^ = 0.725, MAE = 6.065 years, Cohen’s f^2^ = 2.635, N = 466 HCs + augmented data; **Fig. 2A**), consistent with other normative aging models in the field using gait^31,32^ and neuroimaging data^8,9,18,33^. Feature importance (F-scores) indicated that speed (F = 820.958, *p* < 0.001) and double support time (F = 802.458, *p* < 0.001) were the most informative predictors, followed by cadence (F = 208.302, *p* < 0.001), stride length (F = 46.824, *p* < 0.001), and sex (F = 12.803, *p* < 0.001; **Fig. 2A**). The model trained with the TILDA dataset also showed robust performance (*r* = 0.440, *p* < 0.001, R^2^ = 0.19, MAE = 5.62 years, Cohen’s f^2^ = 0.24, N = 1,080 HCs and no augmented data; **Fig. 2D**). In this model, top features included speed-based metrics, followed by stance and cycle times (**Fig. 2D** and **Supplementary Table S3**).

**Figure 2.**
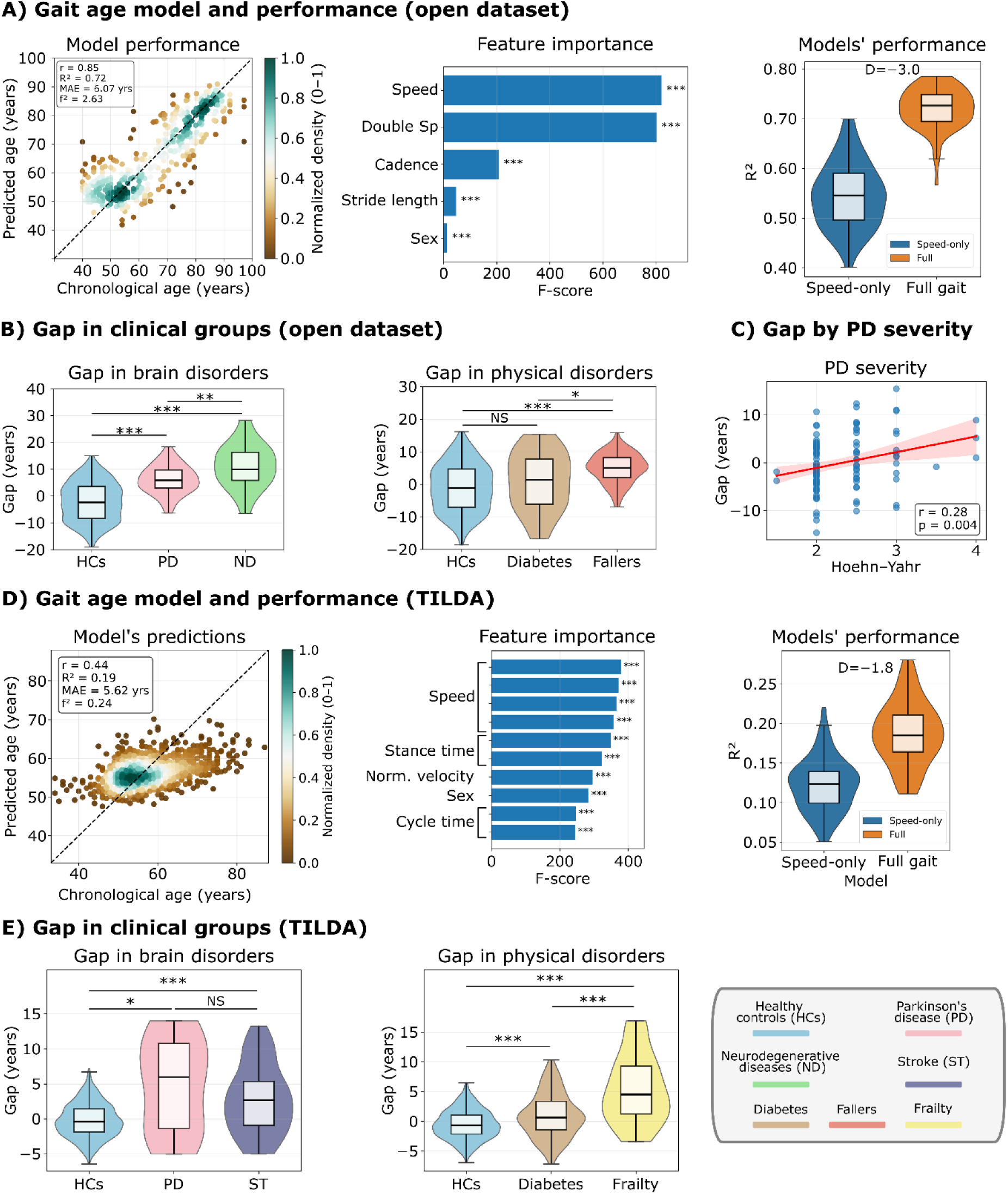
Gait age and gaps. **A)** Clocks from the “open dataset”. The model performance was assessed using Pearson’s correlation and mean absolute error (MAE) between chronological age and predicted age (the “gait age”). Performance values displayed correspond to *r* = 0.85 (*p* < 0.001), R^2^ = 0.72, MAE = 6.07 years, Cohen’s f^2^ = 2.63. No age bias correction was used to report the model performance. Feature importance was measured using F-scores (higher values indicate more informative features). We compared the full gait model performance (four gait metrics plus sex) with the speed-only model (speed plus sex) using the R^2^; box plots were generated from the performance values across pooled folds (15 repetitions). **B)** Gap comparisons in the “open dataset”. The gaps were compared between HCs and different clinical groups: Parkinson’s disease (PD), neurodegenerative diseases (ND, comprising Huntington’s disease and amyotrophic lateral sclerosis), diabetes, and fallers. Groups were compared using ANCOVA, controlling for age and sex. Higher gaps are related to poor general health (accelerated aging). **C)** Correlation between gaps and the Hoehn-Yahr scale for PD stage progression. Gaps were residualized for age and sex (partial correlations), but the Hoehn-Yahr scores were kept untouched. Dots in the scatter plot correspond to participants (including 50% of augmented data). Associations were determined using Pearson’s correlation, with the red line and shaded area representing the regression line and the 95% ordinary least squares confidence interval, respectively. **D)** Clocks from the TILDA dataset, with *r* = 0.44 (*p* < 0.001), R^2^ = 0.19, MAE = 5.62 years, Cohen’s f^2^ = 0.24. The TILDA’s speed-only model consisted of the gait speed alone, like the model in (A). **E)** Gap comparisons in the “TILDA dataset”, where we considered HCs, PD, stroke (ST), diabetes patients, as well as participants with frailty. Outliers were excluded as values beyond ±2 standard deviations from the mean. All group comparisons were made using Welch’s t-test (two-sided). All *p*-values were FDR-corrected. NS: non-significant, *: *p* < 0.05, **: *p* < 0.01, ***: *p* < 0.001. Violin plots were built using the median, the 25th and 75th quartiles, and the data range.

To quantify the added value of full gait patterns beyond walking speed, we compared the full model (four gait metrics + sex) to a speed-only model using R^2^ from folds and repetitions. The full model outperformed the speed-only model in R^2^ value (R^2^ speed-only = 0.545; R^2^ full gait model = 0.720; ΔR^2^ = 0.175, Cohen’s D = 2.965; **Fig. 2A**). We confirmed these results in the TILDA dataset (R^2^ speed-only = 0.122; R^2^ full gait model = 0.188; ΔR^2^ = 0.066, permutation D = 1.841; **Fig. 2D**).

### Gaps showed accelerated aging in clinical conditions

Gaps differed between HCs, PD, and ND after controlling for age and sex (Welch’s F = 107.257, *p* < 0.001; **Fig. 2B**). Pairwise tests showed higher gaps in PD than in HCs (Δgap = 8.126, *t*(190.761) = 12.858, FDR-corrected *p* < 0.001, D = 1.173; **Fig. 2B**) and higher gaps in ND than in HCs (Δgap = 13.237, *t*(30.964) = 8.640, FDR-corrected *p* < 0.001, D = 1.819; **Fig. 2B**). ND conditions also showed higher gaps than PD (Δgap = 5.111, *t*(35.418) = 3.222, FDR-corrected *p* = 0.003, D = 0.845; **Fig. 2B**). Physical conditions (diabetes and fallers) also showed elevated gaps compared to HCs (Welch’s F = 48.335, *p* < 0.001; **Fig. 2B**). Fallers showed higher gaps than HCs (Δgap = 5.991, *t*(166.987) = 9.873, FDR-corrected *p* < 0.001, D = 0.860; **Fig. 2B**), and higher gaps than diabetes (Δgap = 4.145, *t*(41.643) = 2.605, FDR-corrected *p* = 0.019, D = 0.672; **Fig. 2B**), but no statistical differences were found between diabetes and HCs (Δgap = 1.846, *t*(37.607) = 1.192, FDR-corrected *p* = 0.241, D = 0.248; **Fig. 2B**). Results were unchanged when using fully matched tandems by demographics (**Supplementary Fig. S1** and **Table S4**).

To test whether gaps track disease progression in PD, we correlated gaps with Hoehn-Yahr stage (PD only). Higher gaps were associated with a more advanced PD stage (*r* = 0.277, *p* = 0.004, f^2^ = 0.083, N = 106; **Fig. 2C**).

Finally, we validated our results in the TILDA dataset (**Fig. 2E**). Gaps differed between HCs, PD, and ST after controlling for age and sex (Welch’s F = 17.300, *p* < 0.001; **Fig. 2E).** Pairwise tests showed higher gaps in PD than in HCs (Δgap = 5.024, *t*(14.056) = 2.954, FDR-corrected *p* = 0.016, D = 1.976; **Fig. 2E**) and higher gaps in ST than in HCs (Δgap = 3.059, *t*(57.527) = 4.652, FDR-corrected *p* < 0.001, D = 1.164; **Fig. 2E**), but no statistical differences between PD and ST were found (Δgap = 1.965, *t*(18.343) = 1.079, FDR-corrected *p* = 0.294, D = 0.371; **Fig. 2E**). Physical conditions (diabetes and frailty) also showed elevated gaps compared to HCs in the TILDA dataset (Welch’s F = 51.830, *p* < 0.001; **Fig. 2E**). We found higher gaps in diabetes than HCs (Δgap = 1.471, *t*(335.909) = 6.189, FDR-corrected *p* < 0.001, D = 0.530; **Fig. 2E**), and in participants with frailty respect to HCs (Δgap = 5.985, *t*(47.814) = 7.184, FDR-corrected *p* < 0.001, D = 2.215; **Fig. 2E**). Frailty also showed higher gaps than diabetes (Δgap = 4.514, *t*(54.111) = 5.252, FDR-corrected *p* < 0.001, D = 1.109; **Fig. 2E**).

### Gaps are associated with health biomarkers

We next tested whether gaps relate to physical health metrics, including a frailty index (global health deficits), the Short Physical Performance Battery (SPPB; a composite index of physical function, with lower scores indicating worse mobility, balance, and strength), and handgrip strength (muscle strength). All metrics (including gaps) were residualised for age and sex (partial correlations) (**Fig. 3A**). Higher gaps were statistically associated with higher frailty (*r* = 0.174, FDR-corrected *p* = 0.043, N = 147, Cohen’s f^2^ = 0.031), and with worse physical performance as indexed by lower SPPB scores (*r* = −0.279, FDR-corrected *p* = 0.002, N = 147, Cohen’s f^2^ = 0.084) and lower handgrip strength (*r* = −0.310, FDR-corrected *p* < 0.001, N = 143, Cohen’s f^2^ = 0.106).

**Figure 3.**
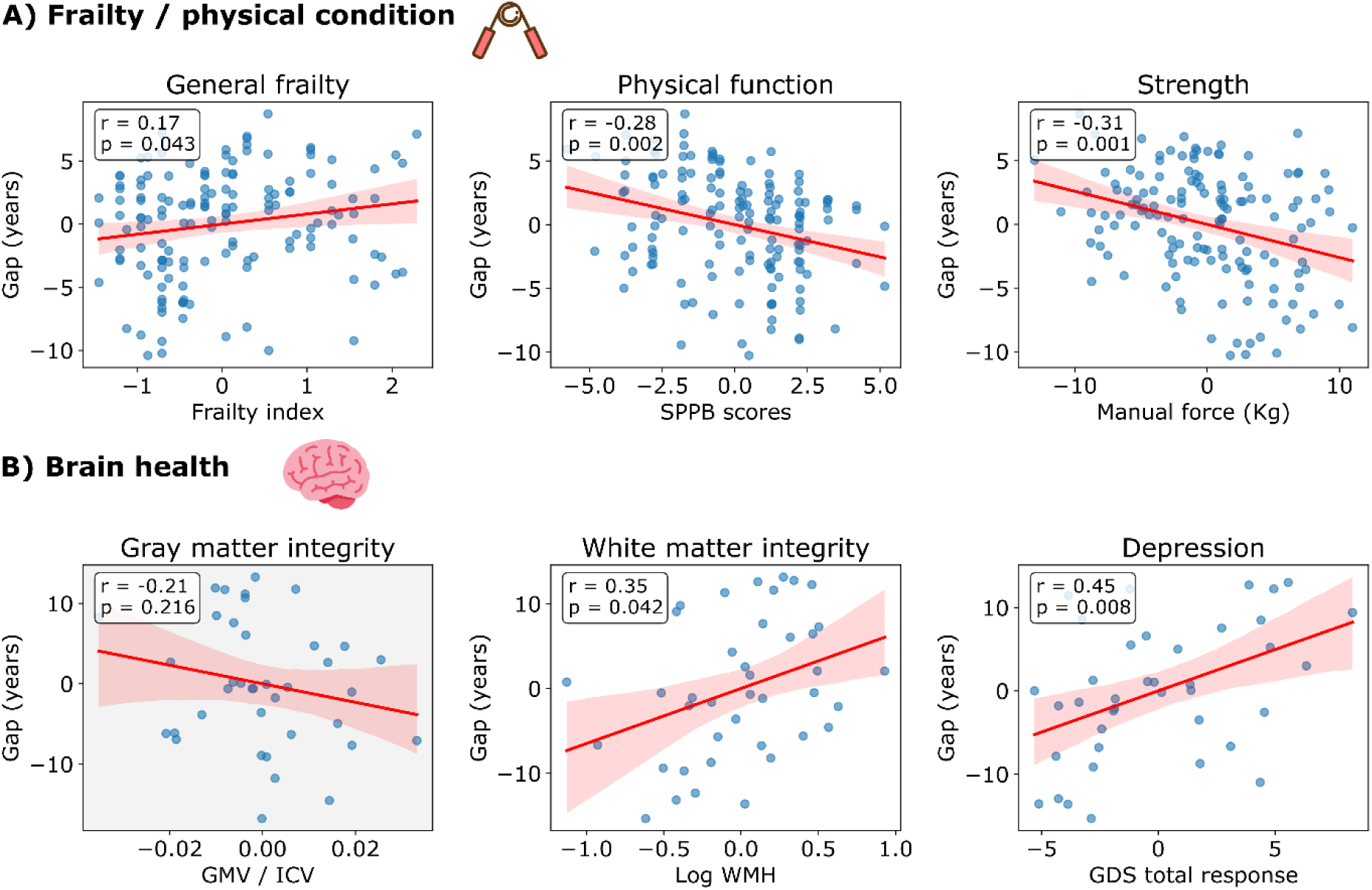
Associations between gaps and clinical biomarkers. **A)** Associations with frailty-related metrics: frailty index, Short Physical Performance Battery (SPPB) score, and handgrip strength (manual force). **B)** Associations with brain health-related metrics: grey matter volume (GMV, normalised by total intracranial volume, ICV), white matter hyperintensities (WMH), and Geriatric Depression Scale (GDS) scores (higher values indicating more depression-like symptoms). All metrics, including the gaps and the different biomarkers, were residualized for age and sex (partial correlations). Outliers were removed from the regression models using Cook’s distance > 4/N (N = sample size). Dots in the scatter plots correspond to participants. Associations were determined using Pearson’s correlation, with the red line and shaded area representing the regression line and the 95% ordinary least squares confidence interval, respectively. All *p*-values were FDR-corrected.

We then linked gaps with brain-related markers, including (i) global grey matter volume (normalised by total intracranial volume; a macrostructural marker of brain tissue volume); (ii) white matter hyperintensities (WMH; higher values indicate greater white matter lesion load); and (iii) Geriatric Depression Scale (GDS; higher values indicate more depression-like symptoms). Using partial correlations (**Fig. 3B**), gaps were not statistically associated with global grey matter volume (*r* = −0.205, FDR-corrected *p* = 0.216, N = 38, Cohen’s f^2^ = 0.044), but were positively statistically associated with WMH burden (*r* = 0.348, FDR-corrected *p* = 0.042, N = 40, Cohen’s f^2^ = 0.138) and GDS scores (*r* = 0.448, FDR-corrected *p* = 0.008, N = 39, Cohen’s f^2^ = 0.251).

### Longitudinal data and lifestyle

In the TILDA dataset, we used Waves 1 (collected between 2009 and 2011) and 3 (collected between 2014 and 2015) to assess longitudinal associations in gait aging. Gaps were weakly but significantly correlated with MRI-based brain age gaps (BAGs) in the TILDA subset using imaging, both when using Wave 1 gait gaps (*r* = 0.113, FDR-corrected *p* = 0.030, f^2^ = 0.013, N = 369) and Wave 3 gait gaps (*r* = 0.165, FDR-corrected *p* = 0.002, f^2^ = 0.028, N = 366; **Fig. 4A**). Gaps were also consistent across time, with Wave 1 gaps predicting Wave 3 gaps (*r* = 0.466, FDR-corrected *p* < 0.001, f^2^ = 0.277, N = 3,077; **Fig. 4A**), indicating that gait aging captures a stable individual signal over time.

**Figure 4.**
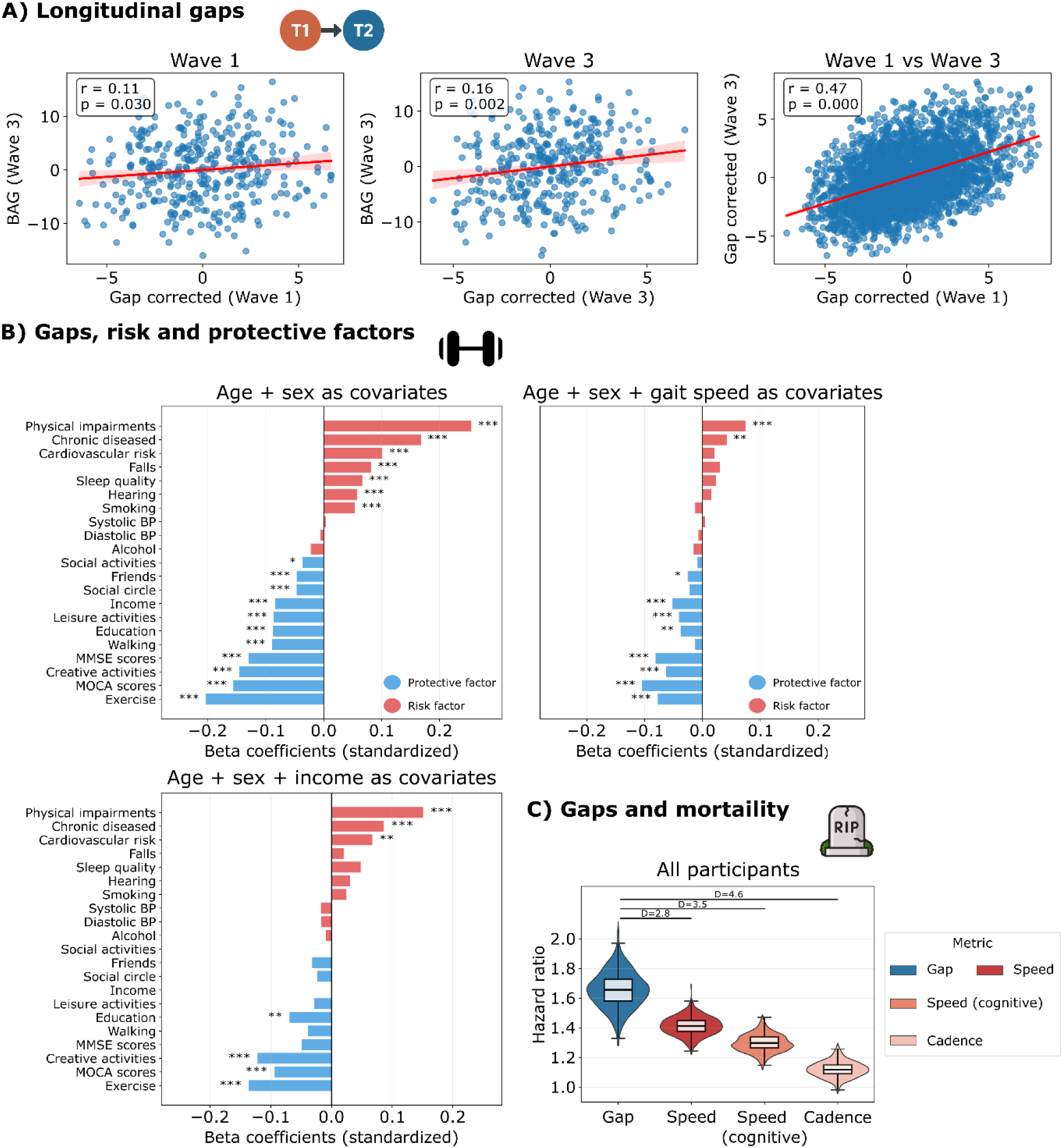
Longitudinal gaps, associated factors, and mortality (TILDA dataset). **A)** Longitudinal analyses of gaps between TILDA Waves 1 and 3, including the association between gaps across Waves and the relationship between corrected gaps at Waves 1 and 3. **B)** Associations between gaps and risk/protective factors. Standardised beta coefficients (β) are shown from regression models adjusted for age and sex, and from models additionally adjusted for gait speed. Positive β values indicate increasing gaps (toward accelerated gait aging), whereas negative β values indicate decreasing gaps (toward delayed gait aging). Bars are color-coded as risk (red) or protective (blue). **C)** Mortality prediction using gaps and individual gait parameters (all metrics z-scored). Hazard ratios (HRs) were estimated using Cox proportional hazards models; HR > 1 indicates increased mortality risk per 1 standard deviation increase in the predictor. Violin plots show the distribution of HR estimates for all participants, healthy controls, and non-healthy controls, with embedded boxplots indicating the median and interquartile range. For association analyses, all metrics, including gaps and biomarkers, were residualized for age and sex (partial correlations). Outliers were removed using Cook’s distance > 4/N (N = sample size). Dots in the scatter plots correspond to participants. Associations were determined using Pearson’s correlation; red lines and shaded areas indicate the regression line and 95% ordinary least squares confidence interval. All p-values were FDR-corrected. NS: non-significant; *: *p* < 0.05, **: *p* < 0.01, ***: *p* < 0.001. Violin plots were built using the median, the 25th and 75th quartiles, and the full data range. Outliers were excluded as values beyond ±2 standard deviations from the mean.

Next, we tested whether gaps were associated with risk and protective factors using Wave 1 TILDA questionnaires and assessment data (**Fig. 4B**). We fitted linear models with standardised variables and gaps, and summarised associations using standardised beta coefficients (β), where the sign indicates the direction of the association. Gaps were associated with diverse risk factors (e.g., cardiovascular health, smoking, chronic disease burden, hearing impairment, falls, and poor sleep) and protective factors (e.g., social and physical activities, number of close friends, creativity/hobbies, and better cognition) (**Fig. 4B; Supplementary Table S5**). These associations remained broadly consistent after additionally adjusting for gait speed and net income (**Fig. 4B; Supplementary Table S5**).

Finally, we evaluated whether gait metrics predicted mortality in TILDA using Cox proportional hazards models (**Fig. 4C**), where hazard ratios (HRs) quantify the increase in mortality risk per 1 standard deviation increase in each gait metric (HR > 1 indicates higher risk). In the full cohort, gaps showed the strongest mortality association (HR, 1.646; 95% CI, 1.417-1.894; FDR-corrected *p* < 0.001), followed by walking speed (HR, 1.409; 95% CI, 1.286-1.528; FDR-corrected *p* < 0.001), dual-task walking speed (HR, 1.301; 95% CI, 1.187-1.423; FDR-corrected *p* < 0.001), and cadence (HR, 1.119; 95% CI, 1.023-1.233; FDR-corrected *p* = 0.018). Consistent with these HR estimates, effect size comparisons indicated that gaps showed stronger mortality associations than walking speed, dual-task walking speed, and cadence (all |D| > 2.7; **Fig. 4C**). Using clinically interpretable contrasts in the original units, a 3-unit higher gap was associated with higher mortality hazard (HR, 1.208; 95% CI, 1.144-1.278; FDR-corrected *p* < 0.001), as was a 0.1 m/s slower walking speed (HR, 1.173; 95% CI, 1.118-1.230; FDR-corrected *p* < 0.001) and a 0.1 m/s slower dual-task walking speed (HR, 1.095; 95% CI, 1.059-1.134; FDR-corrected *p* < 0.001), whereas the association for a 10 steps/min lower cadence did not survive FDR correction (HR, 1.075; 95% CI, 0.980-1.178; FDR-corrected *p* = 0.134).

## Discussion

We examined whether gait dynamics, captured across multiple gait parameters, could provide biologically meaningful measures of aging and how these measures related to clinical, functional, and lifestyle factors in a large-scale sample of 5,681 participants. Gait age clocks showed robust age prediction and outperformed models based on gait speed alone, indicating that gait dynamics, beyond gait speed, might capture the nuances of healthy aging better. Gait age gaps were associated with accelerated aging across neurological and physical conditions and scaled with disease severity (i.e., Parkinson’s disease stages). The gaps were also associated with frailty, physical function, white matter hyperintensities, geriatric depression, and MRI-based brain age gaps. They were additionally linked to diverse risk and protective lifestyle factors (e.g., physical activity, cardiovascular risk, hobbies/creative activities, and cognition), and these associations were broadly consistent after adjustment for net income and gait speed. Gait age gaps also predicted mortality more strongly than any individual gait parameter, including walking speed, paralleling observations from brain and organ aging models in clinical populations. This work provides evidence that gait-based normative modelling may capture a scalable and potentially clinically relevant signal of biological aging with potential utility for risk stratification, monitoring, and personalized medicine.

The main contribution of this work lies in the clinical characterization of the gait age gap. Although this concept is not new in the field of normative modelling of aging, the gait age gap emerges as a promising biomarker with potential real-world applications. Similar to previous work^5,34^, the full gait model outperformed the speed-only model in two independent datasets, supporting the added value of integrating different views of gait dynamics, rather than relying on gait speed alone. The gait age gap derived from this model also showed properties consistent with a clinical biomarker of accelerated aging. In neurological conditions, it reproduced a pattern that closely resembles findings from MRI-based brain clocks, where disorders such as Parkinson’s disease^9,35,36^ Alzheimer’s disease^8,9,18^, and stroke^9^ are associated with older biological/brain age. In our study, this pattern was further supported by the fact that gaps increased with Parkinson’s disease stage, as reported previously using brain clocks in both Parkinson’s disease^35^ and Huntington’s disease^37^, suggesting sensitivity to disease burden and progression. A similar gradient was observed in physical conditions, where frailty and falls showed larger deviations than diabetes. One hypothesis could be that frailty and falls reflect more direct and multisystem functional impairment than diabetes as a diagnosis category, encompassing deficits in balance, variability, and postural control^38^. Therefore, the value of gait age and gaps gets more relevant in conditions where multiple aspects of the gait dynamics are compromised, for example, in Parkinson’s disease, which is characterized by alterations not only in gait speed but also in balance and double support time^4^. Altogether, the results indicate that the gait gap is not only sensitive to neurological dysfunction but also to the severity of broader functional impairment.

The gait age gaps were associated with common clinical biomarkers of physical and brain health, i.e., frailty, strength, white matter integrity, and depression, supporting their potential future use as a health biomarker in aging and disease. The links between gait age gaps and established protective and adverse factors further support their potential utility for risk stratification in clinical settings, even beyond socioeconomic context (net income) and gait speed. This converges with previous work on brain age gaps, where lifestyle^9^, physical activity^21,39^, and enriching experiences^20^ have been associated with accelerated and delayed biological aging. By contrast, the attenuation of some associations after accounting for gait speed suggests that certain risk/protective factors (e.g., cardiovascular risk, falls, smoking) may act mainly through generalized slowing, whereas others relate to broader aspects of gait dynamics (e.g., chronic diseases, social activities, hobbies). Some of the factors showing stronger associations in our analyses map onto domains highlighted by the 2024 Lancet Commission^40^, as modifiable dementia risk factors, including physical inactivity, lower education, social isolation, hypertension, smoking, and hearing loss, which supports the clinical relevance of these correlates. Mortality findings further strengthen the clinical worth of the gait age gap. Gait speed is one of the most established functional predictors of survival in older adults^3,41^, even when compared to more complex multivariate models^3^. In our study, gait age gaps were more informative than gait speed for mortality prediction. The convergence across clinical biomarkers, lifestyle-related factors, and mortality supports gait age gaps as a promising longitudinal and scalable marker for clinical monitoring and stratification.

The results presented here also have direct implications for real-world translation and personalised medicine. Gait speed is already used in clinical practice because it is simple, interpretable, and clinically informative^3,41^. Our results suggest that gait age gaps may retain these practical advantages while summarizing a richer set of gait parameters into a single interpretable score. Improvements in IMU-based gait assessment now make wearable and mobile devices a realistic option for gait quantification beyond specialized laboratory platforms^10,15,42^. In parallel, reliable pipelines for computing gait parameters from wearable sensors are now available^43,44^. This raises the possibility of automated, real-time gait assessment in routine care, reducing dependence on specialist processing while improving efficiency and quality control at the point of testing. In this context, gait age gaps might support low-cost, high-frequency home monitoring, disease tracking, and the early detection of deviations from healthy aging trajectories, including prodromal stages of disease. The present framework may also motivate condition-specific normative models to improve patient stratification. More broadly, the associations with modifiable risk and protective factors suggest potential utility for tailored interventions and preventive strategies. Earlier identification of unhealthy trajectories may help reduce clinical burden and downstream healthcare demand^45^.

A major strength of this study is the scale of the pooled sample, which included more than 5,000 participants across healthy aging and multiple neurological and physical conditions. Another strength is the convergence of the findings across two independent datasets, supporting the robustness of our results across different sets of gait parameters, sample sizes, and machine learning models. In addition, the mortality association observed for slower gait speed was similar in magnitude to that reported previously in an independent cohort: in our sample, a 0.1 m/s slower gait speed was associated with an approximately 17% higher mortality hazard, compared with 14% in Studenski et al. (2011)^3^. The longitudinal follow-up further supports the temporal relevance of the gait age gap and its utility as a predictor of mortality. We also carefully addressed key confounders, including age, sex, income, and gait speed, which increased the specificity of the associations with clinical and lifestyle factors. Finally, the models rely on spatial and temporal gait parameters that are simple to derive and clinically interpretable, which increases their translational potential and supports future explorations using wearable and mobile technologies.

Despite these strengths, several limitations should be considered. First, some important clinical populations were not represented, including Alzheimer’s disease and related dementias. Extending this framework to preclinical and prodromal stages of brain and physical disorders will also be important to determine its value for prevention and early detection. Second, the association between gait age gaps and MRI-based brain age gaps was modest. This may reflect measurement noise and other uncontrolled sources of variability, but it may also indicate that brain and physical aging become partly distinct, although overlapping, trajectories. Prior work has likewise reported modest associations between gait and structural brain measures^2^. Third, the observational nature of the data limits causal inference. Higher gait age gaps may reflect baseline differences in health status rather than ongoing accelerated aging^46^, a question that will require longitudinal data from clinical populations. Another limitation is that some gait measures, particularly variability metrics, can show lower reliability than mean spatiotemporal parameters, especially under shorter or less standardized recording protocols, which may have affected both clock performance and downstream associations^47^. Finally, future versions of the model should be trained and tested in more ethnically and geographically diverse cohorts, as the strong contribution of TILDA may limit the generalizability of the present findings.

Overall, our findings support gait age gaps as an interpretable and scalable marker of biological aging across health and disease. The gaps captured clinically meaningful variation beyond gait speed alone and were linked to disease severity, health biomarkers, lifestyle factors, and mortality risk. These results support the exploration of future implementation of normative gait modelling in personalized medicine, including clinical and in-home monitoring, risk stratification, and wearable-based applications.

## Methods

### Participants and Demographics

We pooled data from different databases (“Open Dataset”, N = 562) and The Irish Longitudinal Study on Ageing cohort (“TILDA dataset”, N = 5,119)^29^, for a total sample of N = 5,681 participants (each participant is a distinct data point). Across the pooled sample, participants included healthy controls (N = 1,391) and non-healthy controls (N = 4,290). The non-healthy controls included the following groups which were used for comparisons, in terms of gap differences: Parkinson’s disease (PD, N = 113), neurodegenerative diseases (Huntington’s disease and amyotrophic lateral sclerosis; ND, N = 24), stroke (ST, N = 60), diabetes (N = 324), fallers (N = 86), and frailty (N = 49). Full demographics are reported in **Table 1**. Participants in the Open Dataset were drawn from five open-source databases (**Supplementary Table S1**)^22–28^; across groups, in the open dataset, we included participants >40 years old.

Inclusion criteria for healthy participants included no diagnosis of chronic diseases, disabilities, impairments, or long-term illness. All participants have obtained approval from the corresponding local ethics committees and collected data in accordance with the Declaration of Helsinki. Additional study-specific inclusion criteria and procedures are described in the original publications.

### Clinical Variables and Biomarkers

We examined physical and brain-health markers (when available) in the source datasets. Brain-related measures included total grey matter volume normalised by total intracranial volume (GMV/ICV) and total white matter hyperintensities (WMH). Mood symptoms were indexed using the Geriatric Depression Scale (GDS). Physical health was captured using a frailty index, the Short Physical Performance Battery (SPPB), and handgrip strength. For PD, disease severity was indexed using the Hoehn-Yahr scale. Not all variables were available for all participants; availability and sample sizes per analysis are reported in **Supplementary Table S1**. In the TILDA dataset, we included the Boyle et al. (2021) pre-computed brain age gaps (BAGs) from grey matter density voxels obtained using structural T1-weighted MRI data^48^.

### Gait Parameters

In the Open Dataset, we used four gait parameters as model inputs: walking speed (m/s), cadence (steps/min), stride length (m), and double support time (%). When available, we used the pre-computed gait variables provided by the original databases. In the database number 1 (**Supplementary Table S1**), we approximated double support time as 100−2× swing phase (%). In datasets where gait timing needed to be derived, gait parameters were computed from force-sensitive resistors embedded in the walkway that recorded foot-ground contact timing for each step. Heel-strike and toe-off events were used to derive stride time and the percentage of time spent in double support, and cadence was computed from mean stride time. Measures were summarised at the participant level. In the TILDA dataset, we used a total of 39 gait parameters available in Waves 1 and 3 of the study, obtained using the GaitRITE pressure-sensing mat (NJ, USA)^49^. These parameters are listed in **Supplementary Table S2.**

### Data Augmentation

To increase the number of observations for training the machine learning model, we generated synthetic samples by interpolation of existing gait features within an augmentation age range of 65-90 years. We used data augmentation for the Open Dataset; its sample size is smaller than TILDA’s. For each augmented sample, we selected a target age uniformly within this range (clipped to the observed age range) and chose a sex in proportion to its frequency in the training data. Within that sex, we identified the two participants whose ages bracketed the target age and created a new gait feature vector by linearly interpolating their feature vectors. We added small random perturbations to avoid duplicates: age jitter (Gaussian noise added to the target age), interpolation-weight jitter (Gaussian noise added to the interpolation weight), and a feature-noise fraction (feature-wise Gaussian noise scaled to each feature’s standard deviation). Augmented samples were appended to the original dataset, generating up to 50% new observations relative to the training set. Augmented data were excluded from group comparisons and associations; they were used just for model training and performance assessment. We did not use data augmentation for the TILDA dataset, as the sample size is large enough for the models’ training.

To the author’s knowledge, there is no established consensus on the optimal synthetic-to-real mixing proportion for normative gait models. We adopted a conservative split: 1/3 of the augmented data relative to the total training data. However, we also compared model performance with and without data augmentation (**Supplementary Fig. S2**).

### Gait Age and Normative Models

Normative models were trained using HCs only. We used N = 311 participants in the Open Dataset (age range 40-97 years, 46% females), and N = 1,080 in the TILDA dataset (age range 34-87 years, 54% females). Gait features were z-scored, and biological sex was included as an additional predictor. We trained support vector regression (SVR) models with an RBF kernel to predict chronological age (the predicted value is referred to as “gait age”). Model selection and performance estimation used nested cross-validation (5 folds, 15 repetitions)^50^. In each outer split, hyperparameters were tuned within the inner loop and evaluated on the held-out outer test fold; final performance values were computed from the outer-loop test predictions pooled across folds and repetitions.

Hyperparameters were tuned by grid search over the SVR regularization parameter (C ∈ [0.1, 1, 10, 40]) and kernel width (gamma ∈ [1×10^−3^, 1×10^−2^, 0.1, 1.0]). We also tuned data-augmentation variability using two settings: (age jitter = 0.5, interpolation-weight jitter = 0.5, feature-noise fraction = 0.5) and (age jitter = 1, interpolation-weight jitter = 1, feature-noise fraction = 1). Model performance was quantified using Pearson’s correlation and mean absolute error (MAE) between chronological age and predicted age. Feature importance was estimated using F-scores. Final model performance was computed from the outer-loop test sets.

To compare the predictive performance of the full gait model and the speed-only model, we analysed the distribution of cross-validated R^2^ values across repeated folds. We compared the distributions in terms of Cohen’s D effect size.

### Gait Age Gaps

From the trained models, we computed gait age gaps as predicted age minus chronological age. Gaps > 0 indicate accelerated aging, and gaps < 0 indicate delayed aging. Gaps were computed within the cross-validation framework and averaged across folds and repetitions. We applied age-bias correction to gaps following prior recommendations. Specifically, within each training split, we fit a linear model predicting gaps from chronological age and applied the fitted parameters to the corresponding test-split gaps to obtain age-bias-corrected gaps. Age-bias correction was used for downstream group and association analyses, but model performance metrics were reported without correction because it can inflate correlations with chronological age (overestimating model performance)^30^.

### Associations with Risk and Protective Factors

We examined associations between gaps and a broad set of risk and protective factors in the TILDA dataset using linear regression models. These factors covered multiple domains, including protective factors (e.g., social and physical activities, cognition) and risk factors (e.g., cardiovascular health and smoking). The full list of variables is provided in **Supplementary Table S5.** For each factor, we fitted two regression models with age-bias-corrected gaps as the outcome. Model 1 included age and sex as covariates. Models 2 and 3 additionally included gait speed or net income to test whether associations were independent of these factors. We limited our analysis to control only by income and speed, considering (i) some factors can be related to socioeconomical status (e.g., hobbies and social activities^51^), (ii) to investigate how informative gaps are in relation to gait speed alone.

Before modelling, gaps and each factor were z-scored, so regression coefficients (β) can be interpreted as standardised effect sizes. This makes interpretability easy, considering all the metrics are on the same scale and, consequently, the β can be compared directly. Sex was included as a categorical covariate. The *p*-values were corrected for multiple testing within each model using false discovery rate (FDR; Benjamini-Hochberg).

### Mortality Prediction and Hazard Ratios

We evaluated the association between gaps and mortality in the TILDA dataset using Cox proportional hazards regression^50,52^. Analyses were performed in three cohorts: all participants, HCs, and non-HCs. Mortality status was defined from the TILDA mortality file (event = death when age at death was available; otherwise, censored). Participants were considered censored when no death was recorded during the follow-up period. In those cases, survival time was calculated up to the last observed age in the dataset. Survival time was defined as follow-up time from baseline age to age at death or censoring. For censored participants, time was censored at the maximum observed age in the dataset. We compared gaps against individual gait metrics, including walking speed, dual-task walking speed, and cadence. Predictors were z-scored before modelling to express hazard ratios (HRs) per 1 standard deviation increase. To keep the direction of effects consistent with gait age gaps (where higher values indicate worse/older gait), the sign of non-gap gait metrics was inverted before analysis (i.e., higher values also reflected worse gait-related risk). For each predictor, Cox models were adjusted for chronological age and sex. HR > 1 indicates increased mortality risk per 1 standard deviation increase in the predictor.

To obtain stable estimates and compare predictors within each cohort, we generated bootstrap distributions of HRs by resampling participants with replacement and refitting the Cox model across 1000 bootstrap repetitions. This procedure yielded one HR distribution per predictor and cohort. We then compared the HR distributions of each gait metric against gaps using Cohen’s D effect size.

### Statistical Analysis

For group comparisons, we used ANCOVA to test group differences in gaps while controlling for age and sex. For post hoc comparisons, we used the residualized gaps (age- and sex-adjusted residuals) and performed pairwise two-sided Welch’s t-tests. Outliers were excluded as values beyond ±2 standard deviations from the mean. For associations, we used Pearson’s correlations and partial correlations (regressed each variable on age and sex and saved the residuals, then correlated *those*); Hoehn-Yahr scores were not adjusted. For regression-based association models, influential observations were removed, excluding cases with Cook’s distance > 4/N (where N is the sample size for the analysis). *P*-values were adjusted for multiple comparisons using the Benjamini-Hochberg false discovery rate (FDR) correction. We report effect sizes using Cohen’s D for group differences (small ≈ 0.2, moderate ≈ 0.5, large ≥ 0.8) and Cohen’s f^2^ for association models (small ≈ 0.02, moderate ≈ 0.15, large ≥ 0.35).

## Data Availability

The five open-source gait databases used in this study are listed in **Supplementary Table S1**, together with their source links. The databases were compiled in a single file available at: https://github.com/carlosmig/Gait_age_and_gaps. TILDA offers access to the datasets for research use through pseudonymised datasets, available publicly and under restricted use. These data were accessed through the TILDA VISTA trusted research environment. Researchers wishing to access the data can find more information and application forms on https://tilda.tcd.ie/data/accessing-data/.

## Code availability

All code used to reproduce the analyses, figures, and results reported in this manuscript is available at: https://github.com/carlosmig/Gait_age_and_gaps. The code used to analyse TILDA data is provided in the same GitHub repository.

## Acknowledgement

This work was supported by the Enterprise Ireland Commercialisation Fund Programme (CF-2024-2450-I) awarded to M.G. C.C.-O., who is supported by Enterprise Ireland (CF-2024-2450-I). All participants who were involved in this research, including all in The Irish Longitudinal Study on Ageing, TILDA, and associated personnel and collaborators. TILDA is funded by Atlantic Philanthropies, the Irish Department of Health and the Health Research Board.

## Competing interests

The authors declare no competing interests.

## Ethics declaration

This study used de-identified data from previously collected cohorts and publicly available datasets. For TILDA, ethical approval for each wave, including Waves 1–3 used here, was obtained from the Faculty of Health Sciences Research Ethics Committee at Trinity College Dublin, and all participants provided written informed consent. For the Health&Gait dataset, participants provided written informed consent, including consent for anonymised data sharing, and the original study was approved by the Review Committee for Research of Cádiz, Spain, in accordance with the Declaration of Helsinki. For the GSTRIDE dataset, the original study was approved by the Ethics Committee for Research with Medicines of Hospital Universitario La Paz (Ref. HULP: PI-4486), and written informed consent was obtained from participants or their relatives when applicable. For the PhysioNet dataset on cerebral perfusion and cognitive decline in type 2 diabetes, the full experimental protocol was approved by the Institutional Review Board of Beth Israel Deaconess Medical Center. For the PhysioNet Gait in Parkinson’s Disease dataset, the original study was approved by the local human studies committee of the Tel Aviv Sourasky Medical Center, and informed written consent was obtained from all participants. All data analysed in the present study were de-identified secondary data, and no new participant recruitment or contact was performed.

## Contributions

C.C.-O. and M.G. conceived the study. C.C.-O., I.S., and M.G. wrote the first draft. M.G. supervised the work and acquired funding. C.C.-O. and F.L. performed the formal analyses. C.C.-O. and F.L. were responsible for data visualization. C.C.-O., F.L., I.S., and M.G. interpreted the data. J.M. resources and data curation. S.K. data curation. All authors reviewed, edited, provided critical comments, and approved the manuscript before submission.

## Supplementary Material

### Supplementary Figures

**Figure S1.**
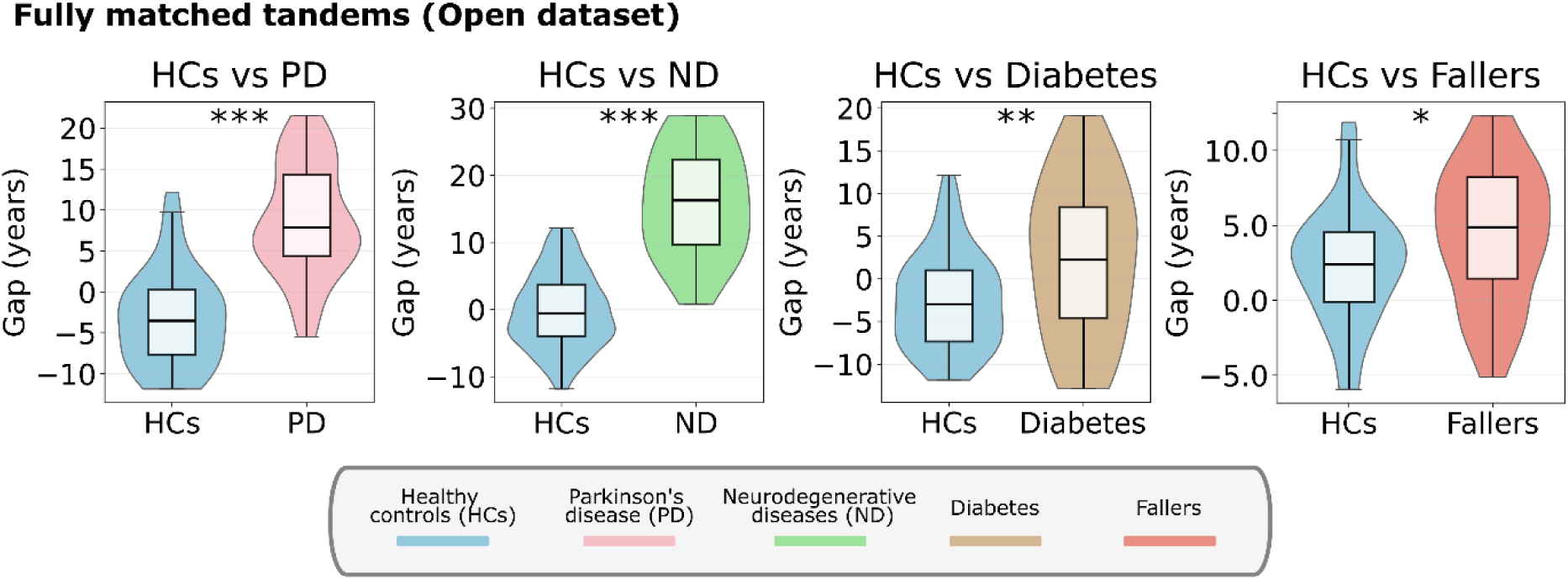
Gaps in fully matched tandems in the “Open dataset”. The gaps were compared between HCs and the different clinical groups in subsamples fully matched by age and sex (ANCOVA was not used in this analysis). Pairwise tests showed higher gaps in patients compared to matched HCs: versus PD (Δgap = 12.04, *t*(93.72) = 9.537, FDR-corrected *p* < 0.001, D = 1.879), ND conditions (Δgap = 16.02, *t*(22.71) = 9.186, FDR-corrected *p* < 0.001, D = 2.777), diabetes (Δgap = 5.063, *t*(46.63) = 3.025, FDR-corrected *p* = 0.005, D = 0.731), and fallers (Δgap = 2.112, *t*(91) = 2.392, FDR-corrected *p* = 0.019, D = 0.492). The violin plots were built using the median, the 25th and 75th percentiles, and the full data range. Outliers were excluded as values beyond ±2 standard deviations from the mean. Group comparisons were made using Welch’s *t*-test. All *p*-values were FDR-corrected. NS: non-significant, *: *p* < 0.05, **: *p* < 0.01, ***: *p* < 0.001.

**Figure S2.**
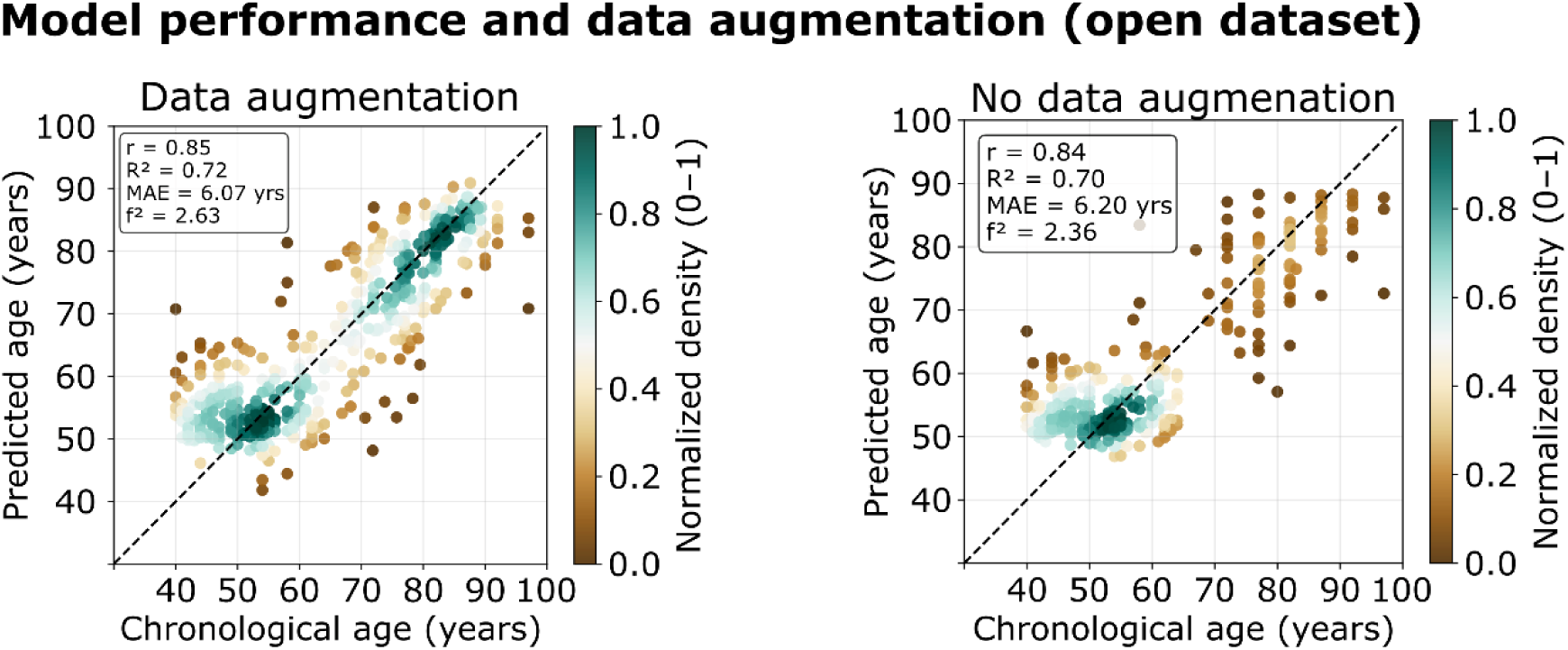
Model performance with and without data augmentation in the open dataset. Predicted age (“gait age”) was plotted against chronological age for the open dataset under two training conditions: with data augmentation and without data augmentation. Each dot represents one sample, and the dot colour indicates the normalized point density (0-1). The dashed diagonal line denotes the identity line, where predicted age equals chronological age. Model performance was assessed using Pearson’s correlation, the R^2^, mean absolute error (MAE), and Cohen’s *f*². In the augmented model, performance values were *r* = 0.85, R² = 0.72, MAE = 6.07 years, and Cohen’s *f*² = 2.63. In the non-augmented model, performance values were *r* = 0.84, R² = 0.70, MAE = 6.20 years, and Cohen’s *f*² = 2.36.

### Supplementary Tables

**Table S1.**
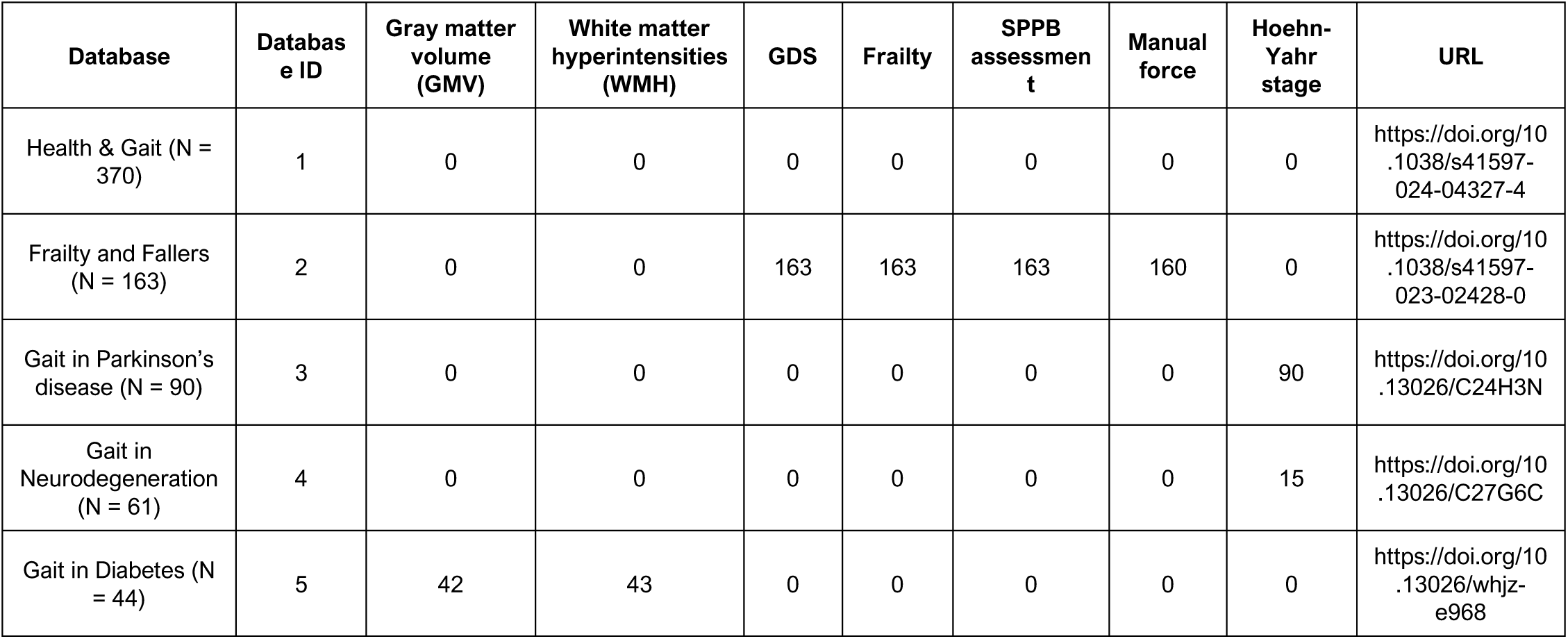
Open dataset variables and sources.

**Table S2.** Gait variables used from the TILDA dataset.

| Domain | Variable | Short description |
| --- | --- | --- |
| Speed / Pace | Normalised walking speed | Walking speed normalised by body size (or leg length, depending on preprocessing). |
| Speed / Pace | Stride velocity (left) | Forward speed estimated from left strides. |
| Speed / Pace | Stride velocity (right) | Forward speed estimated from right strides. |
| Speed / Pace | Walking speed | Usual walking speed during single-task walking. |
| Speed / Pace | Walking speed (cognitive task) | Walking speed during the dual-task / cognitive walking condition. |
| Rhythm / Temporal structure | Cadence | Number of steps per minute. |
| Rhythm / Temporal structure | Gait cycle time (left) | Time to complete one gait cycle with the left leg. |
| Rhythm / Temporal structure | Gait cycle time (right) | Time to complete one gait cycle with the right leg. |
| Spatial parameters | Step length (left) | Distance covered by one left step. |
| Spatial parameters | Step length (right) | Distance covered by one right step. |
| Spatial parameters | Stride length (left) | Distance covered in one full left gait cycle. |
| Spatial parameters | Stride length (right) | Distance covered in one full right gait cycle. |
| Spatial parameters | Step length asymmetry/differential | Difference in step length between left and right sides. |
| Variability (spatial) | Step length variability (left) | Step-to-step variability in left step length (SD). |
| Variability (spatial) | Step length variability (right) | Step-to-step variability in right step length (SD). |
| Variability (spatial) | Stride length variability (left) | Stride-to-stride variability in left stride length (SD). |
| Variability (spatial) | Stride length variability (right) | Stride-to-stride variability in right stride length (SD). |
| Variability (temporal) | Step time variability (left) | Step-to-step variability in left step time (SD). |
| Variability (temporal) | Step time variability (right) | Step-to-step variability in right step time (SD). |
| Variability (temporal) | Stride time variability (left) | Stride-to-stride variability in left stride time (SD). |
| Variability (temporal) | Stride time variability (right) | Stride-to-stride variability in right stride time (SD). |
| Variability (temporal) | Stance time variability (left) | Variability in left stance time (SD). |
| Variability (temporal) | Stance time variability (right) | Variability in right stance time (SD). |
| Variability (temporal) | Swing time variability (left) | Variability in left swing time (SD). |
| Variability (temporal) | Swing time variability (right) | Variability in right swing time (SD). |
| Base of support / stability | Base of support variability (left) | Variability in left base of support (SD). |
| Base of support / stability | Base of support variability (right) | Variability in right base of support (SD). |
| Gait phase percentages | Stance phase (% cycle, left) | Percentage of gait cycle spent in stance (left). |
| Gait phase percentages | Stance phase (% cycle, right) | Percentage of gait cycle spent in stance (right). |
| Gait phase percentages | Swing phase (% cycle, left) | Percentage of gait cycle spent in swing (left). |
| Gait phase percentages | Swing phase (% cycle, right) | Percentage of gait cycle spent in swing (right). |
| Gait phase durations | Stance time (left) | Absolute duration of stance phase (left). |
| Gait phase durations | Stance time (right) | Absolute duration of stance phase (right). |
| Gait phase durations | Swing time (left) | Absolute duration of swing phase (left). |
| Gait phase durations | Swing time (right) | Absolute duration of swing phase (right). |
| Base of support / stability | Base of support (left) | Width of support during left-side stepping/stance. |
| Base of support / stability | Base of support (right) | Width of support during right-side stepping/stance. |
| Foot progression / alignment | Toe-in / toe-out angle (left) | Foot progression angle of the left foot during walking. |
| Foot progression / alignment | Toe-in / toe-out angle (right) | Foot progression angle of the right foot during walking. |

**Table S3.** Ranked feature importance (F-scores) for SVM age-prediction models using gait parameters in the open dataset and TILDA.

| Dataset | Rank | Gait parameter | F-score | p-value |
| --- | --- | --- | --- | --- |
| Open dataset | 1 | Speed | 820.9580 | $p < 0.001$ |
| Open dataset | 2 | Double support time | 802.4580 | $p < 0.001$ |
| Open dataset | 3 | Cadence | 208.3020 | $p < 0.001$ |
| Open dataset | 4 | Stride length | 46.8240 | $p < 0.001$ |
| Open dataset | 5 | Sex | 12.8030 | $p < 0.001$ |
| TILDA | 1 | Stride velocity (right) | 378.5764 | $p < 0.001$ |
| TILDA | 2 | Stride velocity (left) | 371.7087 | $p < 0.001$ |
| TILDA | 3 | Walking speed (cognitive task) | 365.3344 | $p < 0.001$ |
| TILDA | 4 | Walking speed | 357.5718 | $p < 0.001$ |
| TILDA | 5 | Stance time (left) | 348.2409 | $p < 0.001$ |
| TILDA | 6 | Stance time (right) | 322.9694 | $p < 0.001$ |
| TILDA | 7 | Normalised walking speed | 295.8819 | $p < 0.001$ |
| TILDA | 8 | Sex | 284.1746 | $p < 0.001$ |
| TILDA | 9 | Gait cycle time (left) | 247.2376 | $p < 0.001$ |
| TILDA | 10 | Gait cycle time (right) | 244.5902 | $p < 0.001$ |

**Table S4.** Fully matched tandems (Open dataset).

| Demographic variable | HCs (N=35) vs PD (N=56) | HCs (N=267) vs ND (HT+ AL) (N=23) | HCs (N=72) vs Diabetes (N=35) | HCs (N=43) vs Fallers (N=50) |
| --- | --- | --- | --- | --- |
| Sex (F:M)<br>Chi-square test | HCs (1:34)<br>PD (0:56)<br>$p = 0.203$ | HCs (140:127)<br>ND (11:12)<br>$p = 0.671$ | HCs (34:38)<br>Diabetes (14:21)<br>$p = 0.481$ | HCs (42:1)<br>Fallers (50:0)<br>$p = 0.278$ |
| Age (years)<br>Welch's *t*-test | HCs (65.80 ± 7.19)<br>PD (67.52 ± 7.92)<br>$p = 0.289$ | HCs (56.42 ± 11.56)<br>ND (55.22 ± 10.69)<br>$p = 0.610$ | HCs (64.76 ± 7.23)<br>Diabetes (66.77 ± 7.78)<br>$p = 0.205$ | HCs (81.91 ± 4.82)<br>Fallers (82.20 ± 4.51)<br>$p = 0.764$ |

**Table S5.** Associations between gaps and risk/protective factors in the TILDA dataset. We correlated the different factors with the gait age gap, controlling for age and sex, and for age, sex, and speed or income (denoted by * in the Table). The β coefficients capture the direction and magnitude of the relationship.

| Factor | Description | Hypothesis | N | $\beta$ | $p$-FDR | $\beta$<br>(speed)* | $p$-FDR<br>(speed)* | $\beta$<br>(income)* | $p$-FDR<br>(income)* |
| --- | --- | --- | --- | --- | --- | --- | --- | --- | --- |
| Exercise | Participate in sport, exercise (frequency) | Protective factor | 4570 | -0.204 | $p < 0.001$ | -0.078 | $p < 0.001$ | -0.137 | $p < 0.000$ |
| MOCA | Montreal Cognitive Assessment (MOCA) scores | Protective factor | 5103 | -0.156 | $p < 0.001$ | -0.104 | $p < 0.001$ | -0.094 | $p < 0.000$ |
| Hobbies, creative activities | Frequency of engagement | Protective factor | 4536 | -0.146 | $p < 0.001$ | -0.062 | $p < 0.001$ | -0.122 | $p < 0.000$ |
| MMSE | Mini-Mental State Examination (MMSE) scores | Protective factor | 5117 | -0.130 | $p < 0.001$ | -0.081 | $p < 0.001$ | -0.049 | $p = 0.118$ |
| Walking | Number of days (in the last 7 days) on which the participant walked for at least 10 minutes. | Protective factor | 5113 | -0.089 | $p < 0.001$ | -0.013 | $p = 0.268$ | -0.039 | $p = 0.098$ |
| Education | Attend classes, lectures | Protective factor | 4527 | -0.088 | $p < 0.001$ | -0.038 | $p = 0.001$ | -0.069 | $p = 0.001$ |
| Leisure activities | Go to films, plays, concert | Protective factor | 4646 | -0.086 | $p < 0.001$ | -0.040 | $p < 0.001$ | -0.028 | $p = 0.244$ |
| Income | Total household net income | Protective factor | 2564 | -0.084 | $p < 0.001$ | -0.051 | $p < 0.001$ | 0.000 | $p = 1.000$ |
| Social circle | Count of close relatives or friends | Protective factor | 5119 | -0.047 | $p < 0.001$ | -0.022 | $p = 0.058$ | -0.024 | $p = 0.272$ |
| Friends | Number of close friends | Protective factor | 5094 | -0.047 | $p < 0.001$ | -0.026 | $p = 0.031$ | -0.032 | $p = 0.159$ |
| Social activities | Visit, phone family, friends | Protective factor | 4695 | -0.037 | $p = 0.024$ | -0.009 | $p = 0.502$ | 0.000 | $p = 1.000$ |
| Alcohol | Standard drinks a week | Risk factor | 4475 | -0.022 | $p = 0.104$ | -0.016 | $p = 0.171$ | -0.009 | $p = 0.697$ |
| Diastolic BP | Seated diastolic blood pressure (mean) | Risk factor | 5099 | -0.006 | $p = 0.690$ | -0.007 | $p = 0.523$ | -0.017 | $p = 0.459$ |
| Systolic BP | Seated systolic blood pressure (mean) | Risk factor | 5099 | 0.004 | $p = 0.801$ | 0.004 | $p = 0.708$ | -0.017 | $p = 0.459$ |
| Smoking | Number of cigarettes smoked on average per day | Risk factor | 5109 | 0.054 | $p < 0.001$ | -0.013 | $p = 0.268$ | 0.025 | $p = 0.299$ |
| Hearing | Self-rated hearing (negative of) | Risk factor | 5119 | 0.058 | $p < 0.001$ | 0.016 | $p = 0.204$ | 0.031 | $p = 0.178$ |
| Sleep quality | Trouble falling asleep (frequency of) | Risk factor | 5119 | 0.067 | $p < 0.001$ | 0.023 | $p = 0.064$ | 0.048 | $p = 0.051$ |
| Falls | Number of falls in the last year | Risk factor | 5113 | 0.082 | $p < 0.001$ | 0.030 | $p = 0.086$ | 0.021 | $p = 0.357$ |
| Cardiovascular risk | Number of cardiovascular risk factors | Risk factor | 5119 | 0.101 | $p < 0.001$ | 0.021 | $p = 0.092$ | 0.067 | $p = 0.002$ |
| Chronic diseased | Number of chronic diseases | Risk factor | 5119 | 0.168 | $p < 0.001$ | 0.042 | $p = 0.001$ | 0.087 | $p < 0.000$ |
| Physical impairments | Number of physical limitations | Risk factor | 5119 | 0.255 | $p < 0.001$ | 0.074 | $p < 0.001$ | 0.151 | $p < 0.000$ |

## Notes

### Competing Interest Statement

The authors have declared no competing interest.

### Author Declarations

All participants have obtained approval from the corresponding local ethics committees and collected data in accordance with the Declaration of Helsinki. Additional study-specific inclusion criteria and procedures are described in the original publications. The datasets used in the study were individual-level data and any individual-level data had been de-identified before use in this study.

